# Clinical Manifestations and Risk Factors of Pediatric Leptospirosis: A Cross-Sectional Study at a Tertiary Care Center in Northern Vietnam

**DOI:** 10.64898/2026.07.19.26357440

**Authors:** Nguyen Thi Thuy, Tran Thi Thu Huong, Hoang Bao Long, Nguyen Van Lam, Andrew W. Taylor-Robinson

**Affiliations:** Vietnam National Children’s Hospital, Hanoi 115000, Vietnam; College of Health Sciences, VinUniversity, Hanoi 100000, Vietnam; Vinmec Times City International Hospital, Hanoi 116000, Vietnam; Center for Innovations in Health Sciences, VinUniversity, Hanoi 100000, Vietnam; Center for Global Health, Perelman School of Medicine, University of Pennsylvania, Philadelphia, PA 19104, USA

**Keywords:** *Leptospira*, leptospirosis, pediatric, neglected tropical disease, zoonotic infection, cross-sectional study, Vietnam

## Abstract

**Background:** Leptospirosis causes around 1 million cases and 60,000 deaths globally annually, predominantly affecting flood-prone tropical regions. Transmitted through contact with water or soil contaminated by animal urine infected with *Leptospira* spp., this bacterial zoonosis is classified as a high-risk Group B infectious disease in Vietnam. However, epidemiological and clinical data are scarce, particularly in pediatric populations. This study evaluated clinical and laboratory characteristics of leptospirosis cases at the National Children’s Hospital, Hanoi, from January 2023 to August 2025.

**Methodology:** All children admitted with probable or confirmed leptospirosis were enrolled. Clinical and laboratory data were analyzed to characterize disease manifestations and identify risk factors for severe leptospirosis. This was defined by the presence of organ dysfunction, including liver or renal failure, hemorrhage (particularly pulmonary), aseptic meningitis, cardiac arrhythmias, pulmonary insufficiency, or hemodynamic collapse.

**Principal Findings:** Of 84 patients (37 confirmed, 47 probable), the mean age was 9.4 years, with 52.4% male and over half aged ≥ 10 years. Fever was the most common presenting symptom (85.7%); gastrointestinal and renal manifestations were frequent, including oliguria in 26.2% of cases. Key laboratory abnormalities included elevated D-dimer (81.8%), elevated C-reactive protein (75.6%), hypoalbuminemia (74.3%), increased liver enzymes (AST 63.3%, ALT 53.2%), and renal impairment (≈ 46%). Severe disease developed in 48.8% of patients, most frequently as acute kidney injury. Hematuria was independently associated with increased severity (OR = 4.89). Conversely, fever, higher baseline albumin, and longer symptom duration prior to hospitalization were associated with a significantly reduced risk of severe disease.

**Conclusions:** Pediatric leptospirosis in this cohort frequently presented with systemic inflammation and multi-organ involvement, particularly renal impairment. Nearly half the patients developed severe disease. Early recognition of renal signs, especially hematuria, and careful monitoring of albumin levels are critical to identifying children at risk for severe progression and optimizing clinical management.

**AUTHOR SUMMARY:** Leptospirosis is a neglected infectious disease that poses a significant global burden, particularly in tropical countries. Infection spreads from animals to humans via contact with water or soil contaminated by animal urine. While it causes severe illness or even death, information on how it impacts children in Vietnam remains extremely limited.

To fill this gap, this study tracked 84 children treated for leptospirosis in Hanoi between 2023 and 2025. The results revealed that the vast majority experienced a fever alongside damage to multiple organs, especially the kidneys. Nearly half the patients experienced severe illness, with acute renal failure being the most common complication. The presence of red blood cells in the urine was a major warning sign for severe disease progression. Conversely, higher blood albumin levels, fever, and longer illness before hospitalization were each linked to a lower risk of a severe outcome.

Leptospirosis is often misdiagnosed as it has symptoms in common with other tropical fevers. These findings underscore the importance of simple laboratory tests to check for signs of kidney damage and to monitor blood proteins to identify vulnerable children early, thereby enabling timely treatment and thus improving their chances of making a full recovery.

## INTRODUCTION

Leptospirosis is a global zoonotic infectious disease caused by spirochetes of the genus *Leptospira*, which exhibit a unique structural mix of both Gram-positive and Gram-negative characteristics [1]. The bacterium is transmitted from animal reservoirs — particularly rodents — to humans either through direct contact of mucous membranes or abraded skin with infected secretions, or indirectly via water and soil contaminated with infectious urine [2]. While many cases remain mild or asymptomatic, progression to severe illness can trigger Weil’s syndrome, which is characterized by fever, hepatorenal dysfunction, and pulmonary hemorrhage, and can rapidly progress to hemodynamic collapse and death if left untreated [3]. Predominantly distributed in tropical and subtropical zones where warm, humid climates favor pathogen survival [3,4], leptospirosis exerts an annual global burden of around one million clinical cases and 60,000 deaths [1]. Consequently, the WHO classifies it as a neglected tropical disease (NTD), and it is frequently termed a “doubly neglected disease” due to its severe underrepresentation in scientific literature relative to its high public health impact across endemic regions like South and Southeast Asia, Oceania, and Latin America [4–7].

In Vietnam, leptospirosis is classified as a hazardous Group B communicable disease, yet official national surveillance data suffer from profound underreporting; the Vietnam Statistical Yearbook recorded only 0 to 19 cases annually between 2016 and 2020 [8–12]. In stark contrast, localized serological studies reveal widespread, silent community exposure. For instance, Tran *et al.* (2021) detected a 9.5% anti-*Leptospira* IgG seropositivity rate among healthy individuals across the Northern, North Central, and Southern regions [13], while Mai *et al.* (2022) found an 8.3% positivity rate among hospitalized, clinically suspected patients in Thai Binh, Ha Tinh, and Can Tho [14]. This severe epidemiological mismatch highlights critical diagnostic deficiencies and major gaps in national surveillance frameworks.

This surveillance blind spot is exceptionally pronounced in pediatric populations. Although children face high environmental exposure through daily activities like playing, swimming, or wading in floodwaters, childhood leptospirosis is rarely diagnosed. Available data show exposure is common — including a notable 12.8% IgG seroprevalence rate reported among children in southern Vietnamese provinces [15] — but the disease easily masquerades as other endemic infections due to its non-specific presentation. In Thailand, for example, leptospirosis accounted for nearly 19% of non-dengue acute febrile illnesses in children and was frequently misdiagnosed as dengue [16]. This pediatric vulnerability is poised to escalate under the pressure of climate change, as rising average temperatures and severe flooding events prolong the environmental persistence of the bacteria and multiply outbreak risks for recreationally exposed children [17,18].

Despite this growing threat, decades of global and domestic leptospirosis research have focused almost exclusively on adult cohorts, community-level epidemiology, or animal and environmental reservoirs [19,20]. In Vietnam, published literature has largely neglected pediatric populations, leaving the clinical presentation, laboratory profiles, and prognostic drivers of severe childhood leptospirosis virtually unexamined [21,22]. Elucidating these factors is essential to improve clinical diagnostic accuracy, optimize therapeutic pathways, and build a more responsive healthcare infrastructure. In order to address this critical knowledge gap, this study aimed to comprehensively analyze the clinical and laboratory characteristics of pediatric leptospirosis at a major tertiary care center in Northern Vietnam and identify the independent risk factors associated with severe disease progression.

## METHODOLOGY

### Study design and setting

This study was conducted at Vietnam National Children’s Hospital (also known as the Vietnam National Hospital of Pediatrics), a national tertiary pediatric referral hospital in Dong Da District, Hanoi, from January 1, 2023, to August 31, 2025.

### Reference laboratory evaluation for leptospirosis

*Leptospira*-specific IgM antibodies were detected using a commercial ELISA kit (Serion Diagnostics, Würzburg, Germany) according to the manufacturer’s instructions. This assay has a reported a sensitivity ranging from 61% to 100% and a specificity from 82% to over 99% in clinical studies [23–25].

### Case definitions of leptospirosis

Diagnosis of suspected, probable and confirmed cases of leptospirosis was applied as follows:

i. **Suspected cases**: both clinical factors and epidemiologic factors were considered (Table 1) [26,27].
ii. **Probable cases** were defined as suspected cases of leptospirosis that met both of these criteria: the first antibody titer (anti-*Leptospira* IgM ELISA) ≥ 20 IU/ml; AND other causes have been excluded (no evidence of alternative etiology).
iii. **Confirmed cases** were defined as suspected cases of leptospirosis showing serum conversion between anti-*Leptospira* IgM ELISA tests performed on acute- and convalescent-phase sera 7– 14 days apart, which met one of the following criteria: antibodies were not detected in the first serum sample, while the second sample tested positive; OR the first antibody titer was ≥ 20 IU/ml, which the second increased more than 4 times.

**Table 1.** Clinical and epidemiological criteria for suspected leptospirosis.

| Clinical factors | Epidemiologic factors |
| --- | --- |
| Acute febrile illness with headache, myalgia and prostration associated with any other clinical signs including:<br>• Conjunctival suffusion<br>• Renal symptoms: Anuria, proteinuria, or oliguria<br>• Jaundice<br>• Hemoptysis and breathlessness, hemorrhages (in intestines and lungs) | Exposure to contaminated environment within a period of one month:<br>• Water possibly contaminated with *Leptospira* (paddy fields/agricultural fields, domestic sewage, livestock waste, flood water, construction site, river, canal, ditch, recreational activities like rowing, white water rafting, etc.)<br>• Exposure to animals (rodents, livestock, |
| • Meningeal irritation<br>• Cardiac arrhythmia or failure<br>• Skin rash<br>• Gastrointestinal symptoms (nausea, vomiting, abdominal pain, diarrhea)<br>• Arthralgia | • domesticated and wild)<br>• Occupational exposure (farmers, outdoor laborers, fisherman, abattoir worker, veterinarians, military, etc.) |

### Patient recruitment and data collection

All eligible pediatric patients with suspected, probable, or confirmed leptospirosis were enrolled in the study. Patients with confirmed coinfection or insufficient clinical data were excluded. The enrolment process is shown in Figure 1. The study comprised both retrospective and prospective components. For the retrospective component, clinical and laboratory data were extracted from paper-based and electronic medical records using a standardized research case report form and entered into a REDCap database for data management and quality control. Medical records were accessed for research purposes between 16 January 2025 and 31 August 2025. When exposure history or other relevant information was unavailable in the medical records, additional information was obtained through structured telephone interviews with parents or primary caregivers. For the prospective component, participants were recruited between 16 January 2025 and 31 August 2025. Collected data included demographic characteristics, exposure history, clinical manifestations, laboratory and imaging findings, treatment, hospitalization duration, and outcomes.

**Figure 1.**
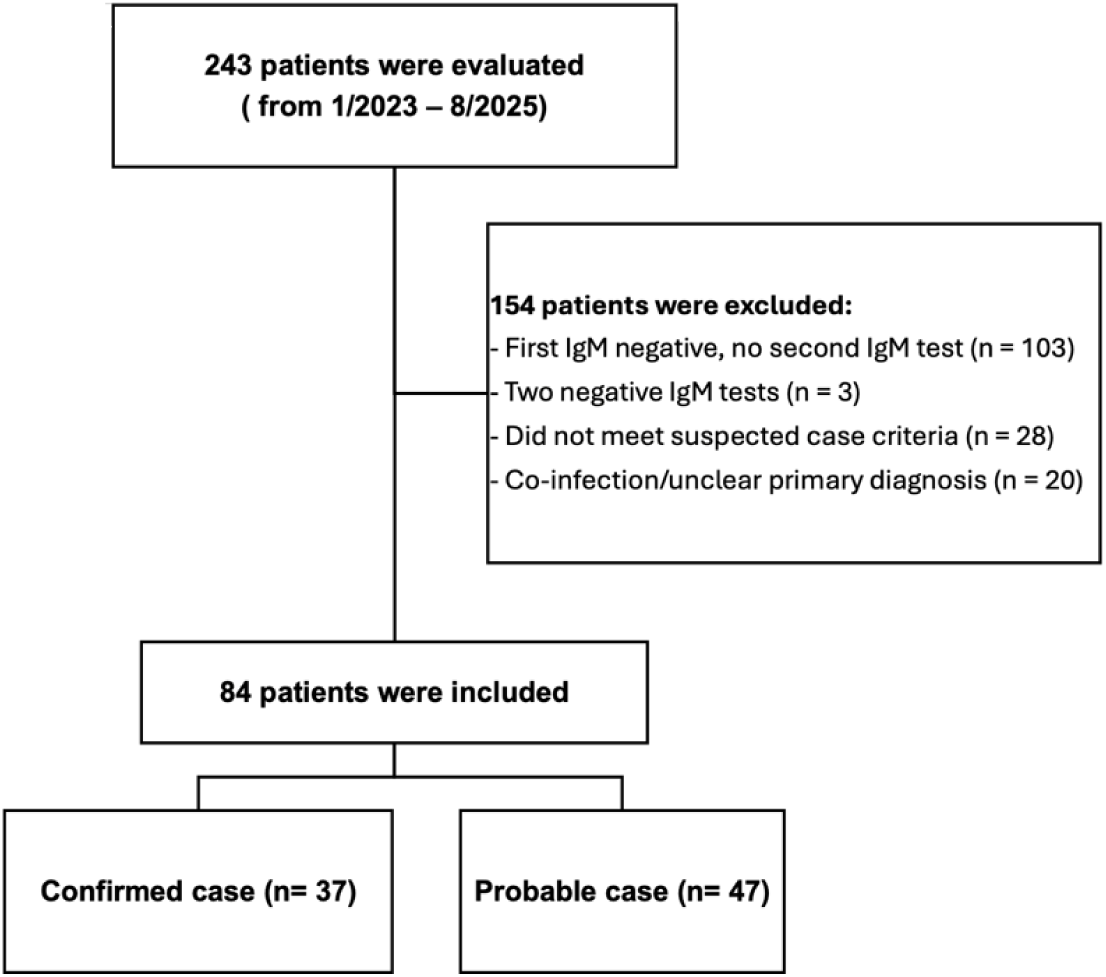
Flow diagram of the enrolment procedure of patients tested for leptospirosis.

### Ethical considerations

The study was approved by the Institutional Review Board of VinUniversity (Approval No. 639/2024/QĐ-VUNI; December 27, 2024) and the Vietnam National Children’s Hospital (Approval No. IRB–VN01037/IRB00011976/FWA00028418; January 15, 2025). For the prospective component, written informed consent was obtained from the parents or legal guardians of all participants before enrolment and data collection. For the retrospective component, verbal informed consent was obtained from parents or primary caregivers before each telephone interview. Study investigators had access to identifiable information only for medical record review and participant follow-up when necessary. All personal identifiers were removed or coded before analysis, and only anonymized data were used for statistical analyses. The study was observational and did not affect patients’ diagnostic or therapeutic management.

### Study variables

The primary outcome was severe leptospirosis, defined by severe organ involvement such as liver failure, renal failure, pulmonary hemorrhage, meningitis, cardiac arrhythmia, respiratory insufficiency, or hemodynamic instability. Independent variables included demographic, epidemiological, clinical, and laboratory characteristics.

### Statistical analysis

Data were managed using REDCap software and analyzed with JASP version 0.95.2. Continuous variables were compared using the independent t-test or Mann–Whitney U test, as appropriate. Categorical variables were analyzed using the Chi-square test or Fisher’s exact test. Univariable and multivariable logistic regression analyses were performed to identify factors associated with severe leptospirosis. Variables with p < 0.20 in univariable analysis or with clinical relevance were included in the multivariable model. A p-value < 0.05 was considered statistically significant.

## RESULTS

### Clinical and laboratory characteristics of children with *Leptospira* infection

#### 1. Demographic and Epidemiological Characteristics of Patients

Among the 84 children with leptospirosis included in the study, males accounted for a slightly higher proportion than females (52.4% vs. 47.6%) (Table 2). The mean age was 9.41 years (range: 1.2–15 years), with more than half of the patients aged ≥ 10 years (51.2%). Most patients were of Kinh ethnicity (the majority ethnic group in Vietnam) (71.4%), and the highest proportion of cases was reported from the Red River Delta region (38.1%), followed by the Northeast region (23.8%) (Figure 1). Environmental and behavioral exposure factors were common: 60.7% of patients reported walking barefoot, 65.5% lived in environments with frequent rat exposure, and 53.5% came from farming or livestock-raising households.

**Figure 1.**
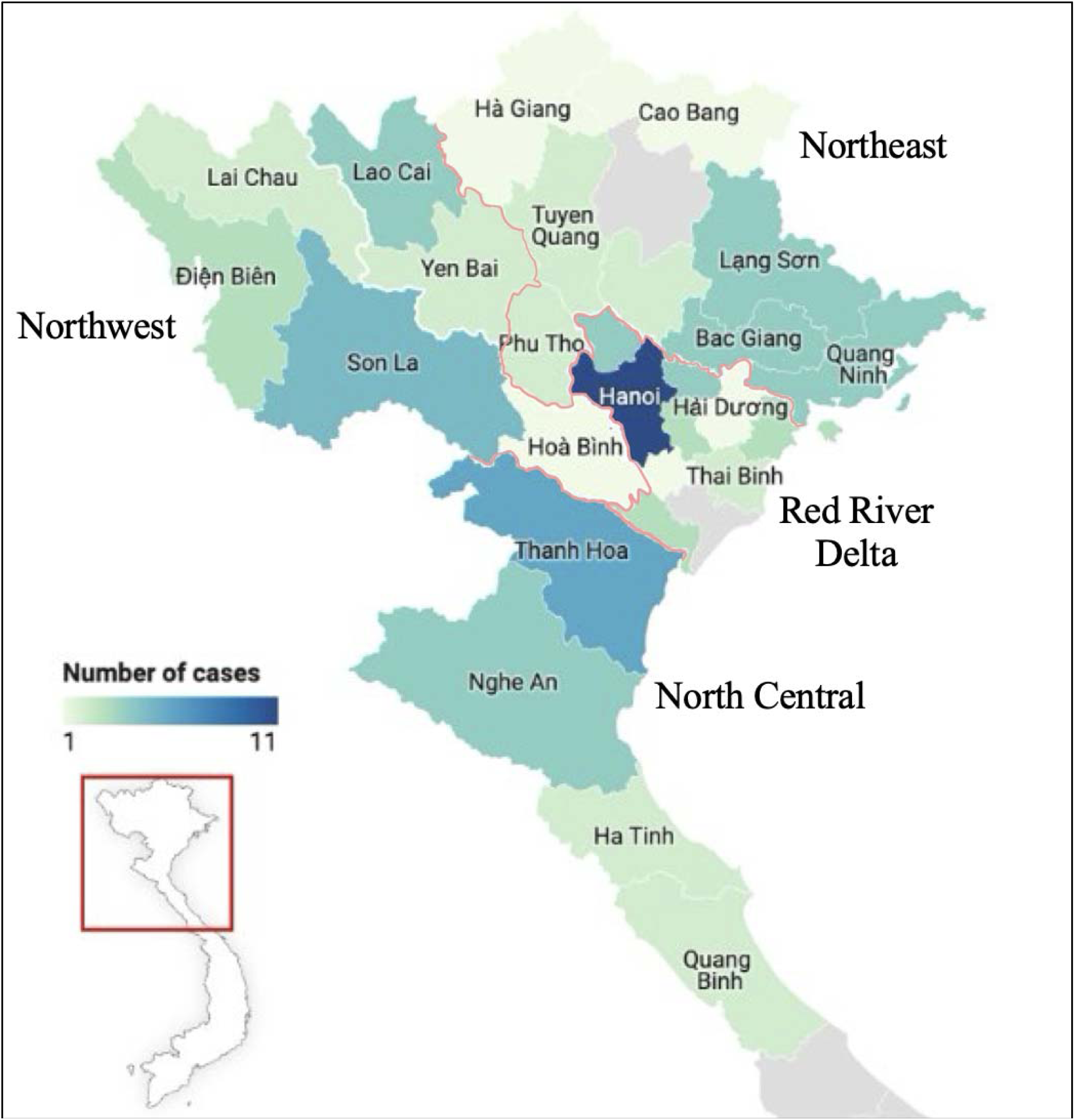
Geographical distribution of pediatric leptospirosis cases (N = 84) by province/city of residence, Vietnam, 2023–2025. Created with app.datawrapper.dep.

**Table 2.**
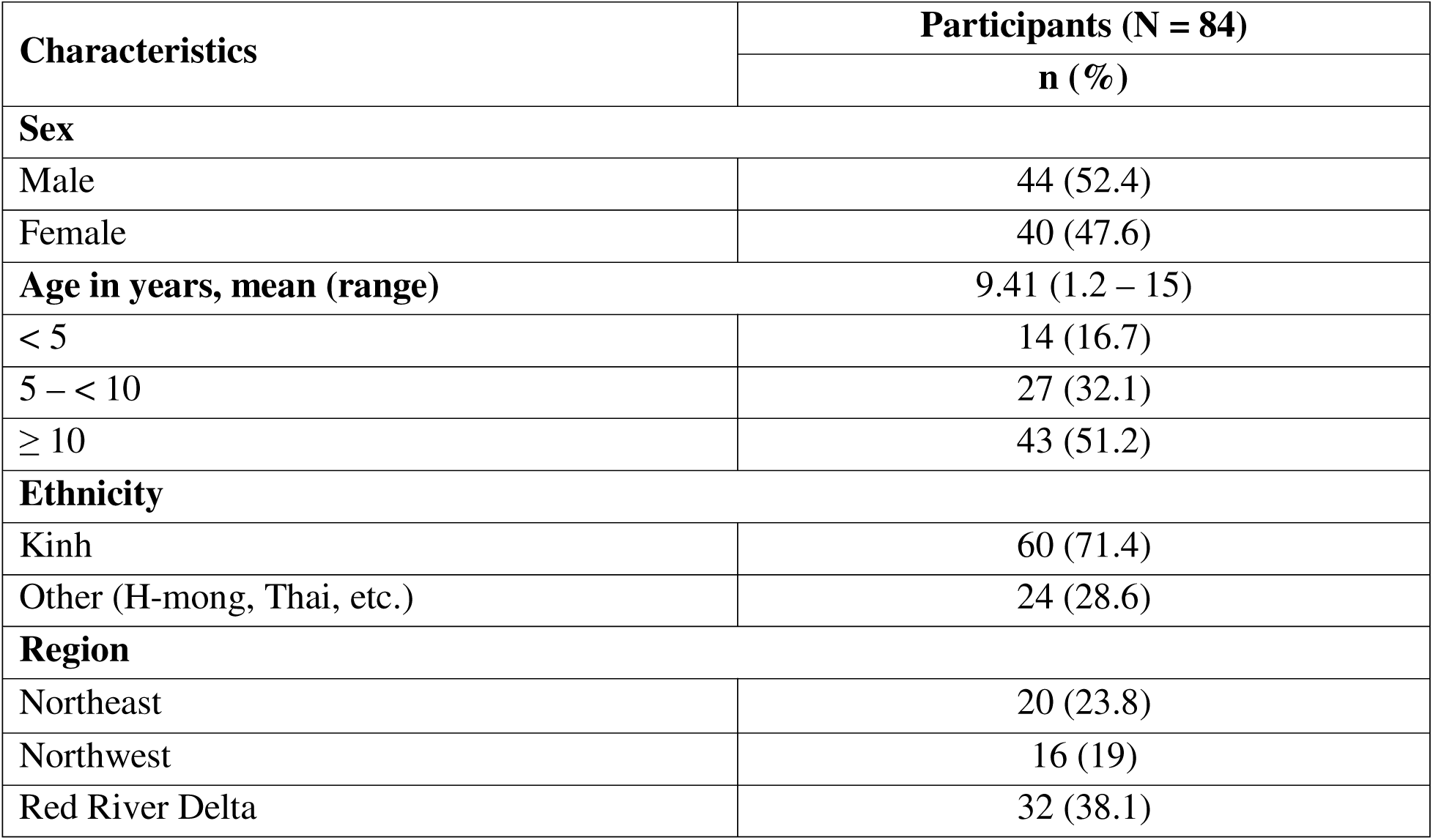
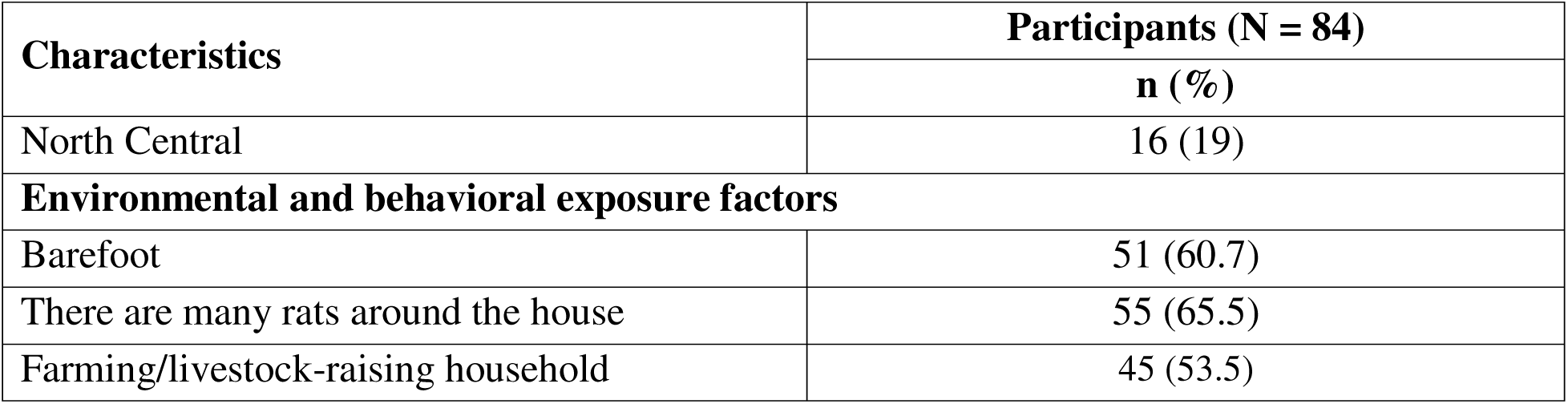
Demographic and epidemiological characteristics of pediatric patients with leptospirosis.

#### 2. Clinical Manifestations

Fever was the most common clinical manifestation (85.7%), followed by fatigue (70.2%) (Table 3). Gastrointestinal symptoms were frequently observed, including abdominal pain (32.1%), vomiting (26.2%), and nausea (25.0%). Respiratory symptoms such as dry cough occurred in 29.8% of patients, while renal manifestations included oliguria (26.2%), edema (14.3%), and anuria (6.0%). The median duration of symptoms before admission was 7 days (IQR: 4–10 days).

**Table 3.** Clinical manifestations of pediatric patients with leptospirosis at presentation.

| Symptoms | Participants<br>N = 84 |
| --- | --- |
|  | n (%) |
| **Systemic** |  |
| Fever | 72 (85.7) |
| Fatigue | 59 (70.2) |
| **Gastrointestinal/Hepatobiliary** |  |
| Abdominal pain | 27 (32.1) |
| Vomiting | 22 (26.2) |
| Nausea | 21 (25.0) |
| Diarrhea | 12 (14.3) |
| Jaundice | 8 (9.5) |
| Melena | 2 (2.4) |
| **Respiratory** |  |
| Dry cough | 25 (29.8) |
| Shortness of breath | 7 (8.3) |
| **Musculoskeletal** |  |
| Muscle aches | 24 (28.6) |
| Joint pain | 2 (2.4) |
| **Renal** |  |
| Oliguria | 22 (26.2) |
| Edema | 12 (14.3) |
| Anuria | 5 (6.0) |
| Hematuria | 4 (4.8) |
| **Neurologic** |  |
| Headache | 12 (14.3) |
|  | n (%) |
| Convulsions | 3 (3.6) |
| **Dermatologic/Mucosal** |  |
| Skin rash | 11 (13.1) |
| Conjunctival suffusion | 3 (3.6) |
| Subcutaneous hemorrhage | 2 (2.4) |
| Duration of symptoms before admission (IQR) | 7 (4–10) |
| Mean fever duration (days) | 5.24 ± 4.36 |

#### 3. Laboratory Findings

Laboratory abnormalities were common and reflected multisystem involvement (Table 4). Elevated inflammatory markers were frequently observed, including CRP (75.6%), ferritin (77.9%), LDH (68.8%), and D-dimer (81.8%). Renal impairment was prominent, with elevated creatinine in 46.9% and elevated urea in 45.8% of patients. Liver involvement was also common, demonstrated by elevated AST (63.3%) and ALT (53.2%). Hypoalbuminemia was seen in 74.3% of cases, while electrolyte disturbances included hypokalemia (39.2%) and hyponatremia (32.4%).

**Table 4.** Abnormal laboratory findings among pediatric patients with leptospirosis.

| Laboratory findings | N | n (%) |
| --- | --- | --- |
| **Complete blood count** |  |  |
| Neutrophilia | 84 | 41 (48.8) |
| Neutropenia | 84 | 13 (15.5) |
| Anemia | 84 | 31 (36.9) |
| Thrombocytopenia | 84 | 14 (16.7) |
| **Inflammatory markers** |  |  |
| Elevated CRP | 82 | 62 (75.6) |
| Elevated Ferritin | 59 | 46 (77.9) |
| Elevated LDH | 48 | 33 (68.8) |
| **Coagulation** |  |  |
| Prolonged PT | 66 | 14 (21.2) |
| Prolonged aPTT | 66 | 6 (9.1) |

| **Laboratory findings** |  | **N** | **n (%)** |
| --- | --- | --- | --- |
| Elevated Fibrinogen |  | 66 | 26 (39.4) |
| Elevated D-dimer |  | 55 | 45 (81.8) |
| **Organ function** |  |  |  |
| Renal function | Elevated urea | 83 | 38 (45.8) |
|  | Elevated creatinine | 83 | 39 (46.9) |
|  | Proteinuria | 56 | 19 (33.9) |
|  | Urine WBC | 84 | 29 (34.5) |
|  | Urine RBC | 84 | 32 (38.1) |
| Liver function | Elevated AST | 79 | 50 (63.3) |
|  | Elevated ALT | 79 | 42 (53.2) |
|  | Elevated direct bilirubin | 39 | 19 (48.7) |
| **Electrolytes** |  |  |  |
| Hypokalemia |  | 74 | 29 (39.2) |
| Hyponatremia |  | 74 | 24 (32.4) |
| **Other** |  |  |  |
| Increased CK |  | 35 | 7 (20) |
| Hypoalbuminemia |  | 74 | 55 (74.3) |
| Decreased C3 |  | 67 | 16 (23.9) |
| Decreased C4 |  | 67 | 5 (7.5) |

Abbreviations: CRP, C-reactive protein; LDH, lactate dehydrogenase; PT, prothrombin time; aPTT, activated partial thromboplastin time; CK, creatine kinase; WBC, white blood cell; RBC, red blood cell; ALT, alanine aminotransferase; AST, aspartate aminotransferase.

Cut-off points and normal reference ranges used to define abnormal laboratory values (e.g., prolonged, elevated, increased, or decreased) were based on the Vietnam National Children’s Hospital Reference Handbook (Version 3, July 20, 2021), an internal institutional laboratory reference document; relevant reference ranges are provided in the Appendix supplementary tables.

#### 4. Severe Leptospirosis and its Complications

Among the 41 patients with severe leptospirosis, most presented with a single severe condition (70.7%), while 22.0% had two severe conditions, and a smaller proportion developed three (2.4%) or four severe conditions (4.9%) (Table 5). Acute kidney injury was the most common severe complication, affecting 63.4% of severe cases, followed by acute liver failure (24.4%) and sepsis (22.0%). Other severe manifestations included circulatory failure (12.2%), coagulation disorders (12.2%), septic shock (4.9%), and respiratory failure (2.4%).

**Table 5.**
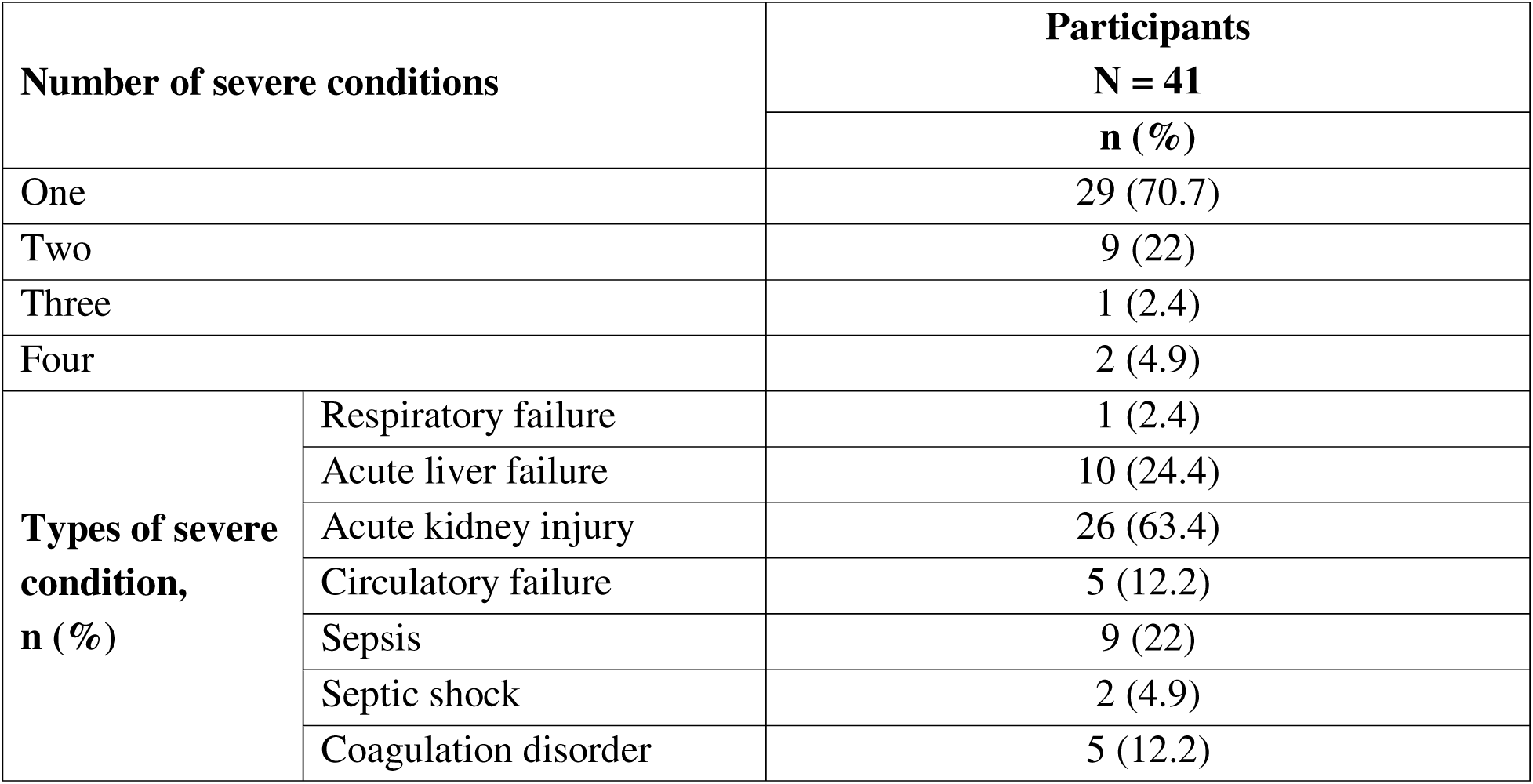
Classification of the number and types of severe conditions.

#### 5. Factors Associated with Severe Leptospirosis

Several clinical features were significantly associated with severe leptospirosis (Table 6). Patients with severe disease more commonly presented with shortness of breath (OR 7.2, p = 0.041), oliguria (OR 7.63, p < 0.001), edema (OR 6.61, p = 0.010), anuria (p = 0.018), and hematuria detected by urine RBC positivity (OR 4.83, p < 0.001). Diarrhea also tended to be more frequent in severe cases (OR 3.75, p = 0.050). These findings suggest that respiratory and renal manifestations were strongly associated with severe pediatric leptospirosis.

**Table 6.** Comparison of clinical features between severe and non-severe leptospirosis.

| Symptom | Non-severe, n (%) | Severe, n (%) | OR (95% CI) | p |
| --- | --- | --- | --- | --- |
| Fever | 42 (97.6) | 30 (73.2) | 0.065 (0.008–0.53) | 0.001 |
| Abdominal pain | 13 (30.2) | 14 (34.1) | 1.197 (0.478–2.992) | 0.701 |
| Vomiting | 9 (21.0) | 13 (31.7) | 1.754 (0.654–4.702) | 0.261 |
| Diarrhea | 3 (7.0) | 9 (22.0) | 3.75 (0.937–15.009) | 0.050 |
| Shortness of breath | 1 (2.3) | 6 (14.6) | 7.2 (0.827–62.683) | 0.041 |
| Oliguria | 4 (9.3) | 18 (43.9) | 7.63 (2.299–25.327) | < 0.001 |
| Edema | 2 (4.7) | 10 (24.4) | 6.61 (1.351–32.371) | 0.010 |
| Anuria | 0 (0) | 5 (12.2) | 13.11 (0.701–254.094) | 0.018 |
| Urine RBC | 9 (21.0) | 23 (56.1) | 4.827 (1.85–12.598) | < 0.001 |

In our study, the severe group had higher ferritin than the non-severe group (p = 0.01). Urea and creatinine were significantly higher in the severe group than in the non-severe group (p < 0.001). Albumin, C3 and C4 were significantly lower in the severe group than in the non-severe group (p < 0.001). CRP, LDH, AST, ALT, direct bilirubin and CK were higher in the severe group than in the non-severe group but did not reach statistical significance (Table 7).

**Table 7.** Blood biochemical characteristics on admission between severe and non-severe leptospirosis.

| Blood biochemical analytes | Non-severe, (n) | Non-severe | Severe, (n) | Severe | p |
| --- | --- | --- | --- | --- | --- |
| **Inflammatory markers** |  |  |  |  |  |
| CRP (mg/L) | 42 | 21.26 | 41 | 36.25 | 0.213* |
| (IQR) |  | (5.34 – 81.51) |  | (12.72– 113) |  |
| Ferritin (ng/mL) | 34 | 243.1 | 25 | 633.6 | **0.01\*** |
| (IQR) |  | (131.3 – 514.8) |  | (313 – 1199) |  |
| LDH (U/L) | 36 | 286 | 30 | 350.4 | 0.064* |
| (IQR) |  | (213.8 – 406.8) |  | (251.8 – 690.5) |  |
| **Organ function** |  |  |  |  |  |
| Ure (mmol/L) | 42 | 3.75 | 41 | 12.1 | < **0.001\*** |
| (IQR) |  | (3.1 – 4.96) |  | (6.16 – 29.2) |  |
| Creatinin (micromol/L) | 42 | 38.5 | 41 | 235.2 | < **0.001\*** |
| (IQR) |  | (31.75 – 50.15) |  | (40.2 – 440.8) |  |
| AST (U/L) | 42 | 46.05 | 37 | 74.5 | 0.064* |
| (IQR) |  | (26.62 – 102.8) |  | (26.1 – 186.8) |  |
| ALT (U/L) | 42 | 34.5 | 37 | 62.2 | 0.473* |
| (IQR) |  | (13.63 – 110.9) |  | (15.2 – 142.2) |  |
| Direct Bilirubin (micromol/L) | 10 | 1.75 | 29 | 5.1 | 0.326* |
| (IQR) |  | (0.68 – 12.2) |  | (1.2 – 33.6) |  |
| Albumin (g/L) | 36 | 37.124 ± 5.363 | 38 | 30.487 ± 5.766 | < **0.001**<sup>^</sup> |
| ( Mean ± SD) |  |  |  |  |  |
| CK (U/L) | 17 | 41.7 | 18 | 107.8 | 0.056* |
| (IQR) |  | (29 – 69.8) |  | (37.4 – 418) |  |
| Electrolytes |  |  |  |  |  |
| Na (mmol/L) | 34 | 135.897 ± 2.584 | 40 | 134.305 ± 4.694 | 0.07** |
| (Mean ± SD) |  |  |  |  |  |
| K (mmol/L) | 34 | 3.71 | 40 | 3.66 | 0.811* |
| (IQR) |  | (3.42 – 3.97) |  | (3.32 – 4.39) |  |
| Complement |  |  |  |  |  |
| C3 (g/L) | 34 | 1.359 ± 0.356 | 33 | 0.922 ± 0.385 | < **0.001**<sup>^</sup> |
| (Mean ± SD) |  |  |  |  |  |
| C4 (g/L) | 34 | 0.343 ± 0.123 | 33 | 0.223 ± 0.104 | < **0.001**<sup>^</sup> |
| (Mean ± SD) |  |  |  |  |  |

**Note:** \*Mann–Whitney U test; \*\* Welch’s *t*-test; <sup>^</sup> Student’s *t*-test; statistically significant p values shown in bold.
Abbreviations: CRP, C-reactive protein; LDH, lactate dehydrogenase; PT, prothrombin time; aPTT, activated partial thromboplastin time; CK, creatine kinase; WBC, white blood cell; RBC, red blood cell; ALT, alanine aminotransferase; AST, aspartate aminotransferase.

Four factors were significantly associated with disease severity (p < 0.05): urine RBC was the primary risk factor (OR = 4.89), whereas albumin, fever at admission, and pre-admission symptom duration were found to be protective factors against severe outcomes (Table 8).

**Table 8.**
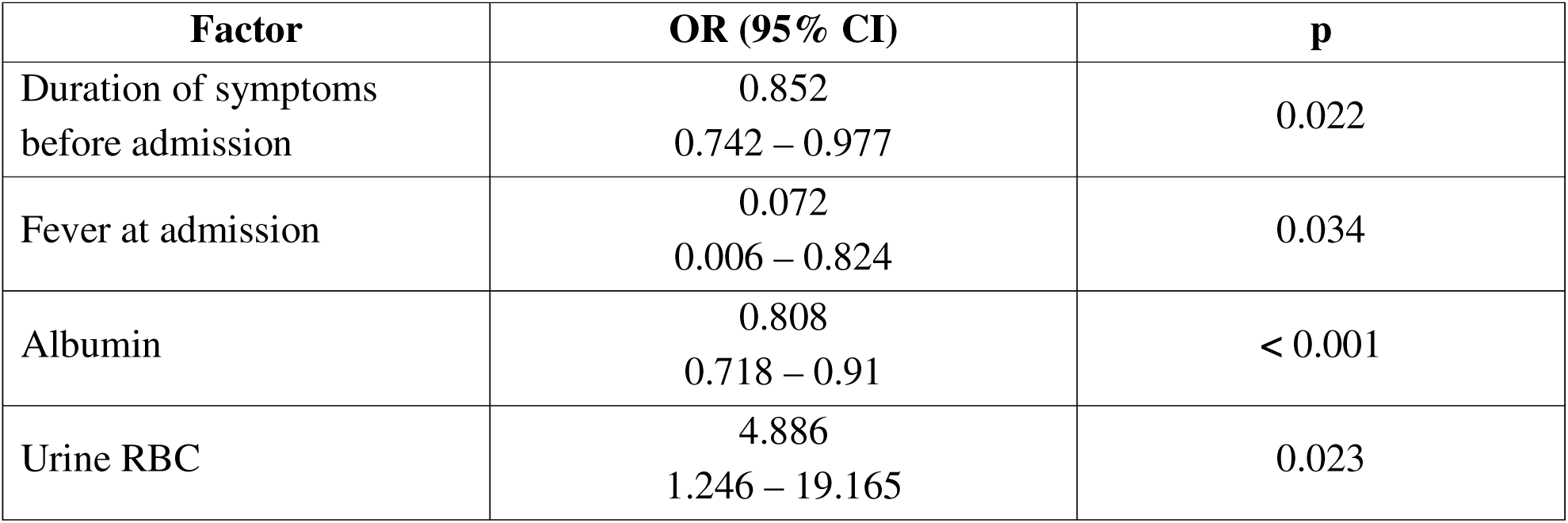
Multivariable logistic regression analysis of factors associated with severe leptospirosis.

| Factor | OR (95% CI) | p |
| --- | --- | --- |
| Duration of symptoms before admission | 0.852<br>0.742 – 0.977 | 0.022 |
| Fever at admission | 0.072<br>0.006 – 0.824 | 0.034 |
| Albumin | 0.808<br>0.718 – 0.91 | < 0.001 |
| Urine RBC | 4.886<br>1.246 – 19.165 | 0.023 |

## DISCUSSION

This cross-sectional study of 84 patients at the Vietnam National Children’s Hospital provides a comprehensive evaluation of pediatric leptospirosis in Northern Vietnam, revealing a substantial burden where nearly half (48.8%) of hospitalized cases progressed to severe disease. To navigate the well-documented challenges of confirming early-phase infections in resource-limited settings, our analysis pooled laboratory-confirmed and probable cases. Importantly, no statistically significant differences in demographic profiles, clinical severity, or baseline laboratory markers were observed between these two sub-cohorts. This homogeneity validates the combined analysis, expanding statistical power and capturing a more representative epidemiological footprint of the disease in line with endemic-region tracking recommendations.

Epidemiologically, our cohort demonstrated a slight male predominance (1.1:1 ratio) and a mean age of 9.4 years, with the majority of patients aged 10 years or older. This distribution mirrors regional patterns where older children, particularly boys, incur higher risks due to cumulative exposure during outdoor recreation, swimming, and agricultural tasks [14,15,28–30]. Geographically, cases were concentrated among children of Kinh ethnicity living within the humid, flood-prone, and river-dense landscapes of the Red River Delta and Northeast regions. The prominence of specific exposure behaviors — namely, walking barefoot, contact with livestock, and residing in rat-infested environments — reaffirms that traditional domestic and agricultural zoonotic reservoirs remain the primary vectors for transmission across Southeast Asia [14,15,25,31–34].

Clinically, most pediatric leptospirosis cases presented with fever and fatigue, with fever lasting a mean of 5 days. Gastrointestinal symptoms were common, whereas jaundice was uncommon, reflecting milder hepatic involvement than in adults [4,28,35–39]. Respiratory manifestations, mainly cough and dyspnea, were observed in a minority of patients and were generally mild. Renal involvement, including oliguria and anuria, occurred infrequently, suggesting that most cases were mild to moderate in severity. Conjunctival suffusion, hemorrhage, and arthralgia were rare but highlighted the broad clinical spectrum of pediatric leptospirosis [28,29,40,41]. The median time from symptom onset to hospital admission was 7 days, consistent with the acute septicemic phase when febrile symptoms are most prominent [4,20,42]. Laboratory profiles revealed intense multisystem inflammation and coagulation activation — characterized by elevated CRP, ferritin, IL-6, fibrinogen, and D-dimer — alongside frequent but moderate hepatic and renal impairment [43–45]. Notably, the pediatric cohort exhibited a distinct, generally milder clinical phenotype than historically documented in adults. Severe adult characteristics, such as deep jaundice, severe thrombocytopenia, overt respiratory failure, and catastrophic pulmonary hemorrhage, were rare, indicating less pronounced pulmonary and hepatic tissue vulnerability in children [4,28,35–39,46].

Our multivariate logistic regression model identified four factors independently associated with disease severity: hematuria (urine RBC), serum albumin, symptom duration prior to admission, and the presence of fever at admission. Notably, microscopic hematuria emerged as the strongest independent risk factor (OR = 4.886, 95% CI: 1.246 – 19.165), yielding a nearly fivefold escalation in severe risk. This correlates precisely with the underlying pathophysiology of *Leptospira* infection, where spirochetes directly invade renal tubules to induce acute interstitial nephritis, driving acute kidney injury (AKI) — the most pervasive severe complication in our cohort. Consequently, the detection of red blood cells on routine urinalysis serves as an invaluable, accessible early warning sign of underlying renal parenchymal damage or subclinical coagulation disorders, mandating immediate and rigorous renal monitoring.

Conversely, serum albumin functioned as a powerful inverse marker of severity. Hypoalbuminemia has been systematically linked to poor outcomes in both leptospirosis and broader critical care settings [47,48]. Rather than simply reflecting baseline nutritional status, a drop in serum albumin serves as a clinical proxy for profound vascular barrier dysfunction, capillary leak, and accelerated systemic inflammation. As a negative acute-phase protein, albumin synthesis is down-regulated during acute stress while concurrent inflammatory cascades increase systemic endothelial permeability [49]. This shifts albumin from the intravascular compartment into the interstitium, a phenomenon frequently observed in severe sepsis cohorts requiring aggressive vasopressor support [48]. Our findings validate its utility as a reliable reflective marker, whereby severe hypoalbuminemia mirrors extensive systemic vascular damage and multi-organ involvement.

A particularly interesting finding from our regression model was the protective association of prolonged symptom duration prior to admission (OR = 0.852, p = 0.022). This inverse relationship indicates that children with a longer course of pre-hospital symptoms experienced a more insidious, lower-grade infection, whereas severe cases manifested with alarming, high-acuity symptoms from the very outset, prompting families to seek immediate tertiary care. This pattern is consistent with historical observations by Sharp et al. (2016), who noted that fatal leptospirosis cases often present early with rapid, distressing manifestations such as jaundice, edema, hemoptysis, and seizures [50]. Similarly, Parra Barrera et al. (2023) identified high-severity clinical clusters characterized by early-onset dyspnea (OR = 5.54), tachycardia (OR = 9.69), and rash (OR = 10.25) [41]. Mechanistically, as highlighted by Rajapakse et al. (2025), severe clinical trajectories are dictated by a volatile interplay of host genetics, pathogen virulence, and an early, dysregulated immune response akin to a cytokine storm [51]. This hyper-inflammatory state is often compounded by high initial leptospiremia and elevated biomarkers such as pNGAL and IL-6, which truncate the incubation-to-crisis timeline and force rapid hospitalization [52].

Equally striking was the finding that the presence of fever at admission was independently associated with a reduced risk of severe progression (OR = 0.053, p = 0.002), framing an afebrile presentation as a strong predictor of severe disease. Several distinct mechanisms explain this clinical paradox. First, in cases of fulminant sepsis or profound septic shock, severe homeostatic collapse can blunt typical febrile pathways or induce hypothermia, indicating an exhausted or impaired host immune response associated with high mortality and widespread multi-organ injury [53,54]. Second, leptospirosis classically follows a biphasic course; patients presenting later in the disease timeline may have already transitioned out of the initial leptospiremic febrile phase and into the immune or complication phase, often masked by prior outpatient antibiotic or antipyretic administration. Finally, contemporary clinical literature confirms that rapidly progressive, afebrile phenotypes frequently occur during acute renal and multi-organ failure [55,56]. Taken together, these data demonstrate that while an active fever reflects an intact, protective immune response, its absence on admission must not rule out leptospirosis, but should instead alert clinicians to an advanced, suppressed, or rapidly accelerating clinical course.

## LIMITATIONS

This study has several limitations. First, its retrospective, cross-sectional nature relies on medical records and parental recall, introducing potential information bias and underestimating subjective symptoms like myalgia or headache. Second, because it was conducted at a single tertiary referral center, selection bias likely overrepresented severe cases (48.8%), limiting generalizability to the broader pediatric community. Third, the small sample size (N = 84) reduces statistical power for analyzing rare clinical presentations or conducting robust subgroup analyses, such as evaluating ethnic minority disparities. Fourth, while combining confirmed and probable cases was methodologically necessary to optimize statistical power, it may limit direct comparisons with global studies relying strictly on MAT or PCR confirmation. Finally, the study lacks serovar characterization to link clinical phenotypes to specific *Leptospira* strains, while the qualitative exposure data lack the precise environmental or rodent reservoir identification required to design highly targeted epidemiological interventions.

## CONCLUSIONS

Pediatric leptospirosis in Northern Vietnam frequently presents as a severe, multi-system inflammatory disease with a high propensity for acute kidney injury, contrasting with the classical multi-organ Weil’s syndrome seen in adults. Our study demonstrates that clinicians do not require cost-prohibitive diagnostic infrastructure to identify high-risk patients; instead, independent risk factors derived from routine, accessible tests — specifically the presence of hematuria, severe hypoalbuminemia, early high-acuity symptom onset, and an afebrile presentation on admission — provide a robust framework for early risk stratification. Incorporating these simple indicators into pediatric triage algorithms will enable timely clinical interventions, optimize fluid and antibiotic management, and ultimately improve survival outcomes for vulnerable children in endemic regions.

Our finding that severe cases were disproportionately concentrated among ethnic minority children and those living in rural or mountainous areas carries significant weight for healthcare governance, signaling potential gaps in localized disease prevention, early detection networks, and primary care access. Addressing these geographic and demographic disparities in Vietnam requires equity-focused policy interventions, including strengthening primary care infrastructure in remote regions, securing timely referral pathways to tertiary centers, and overcoming cultural, linguistic, and financial barriers to care. Ultimately, mitigating the burden of pediatric leptospirosis demands coupling precise clinical risk stratification with targeted health system strengthening.

## Data Availability

This study involved de-identified sensitive patient information that, due to local regulations and ethical frameworks, cannot be publicly shared. Limited access to data sets is available upon reasonable request to the Institutional Review Board of Vietnam National Children's Hospital (contact).

## DECLARATIONS

### Ethics approval

This study was approved by the Institutional Review Boards of VinUniversity (639/2024/QĐ-VUNI, dated December 27, 2024) and the Vietnam National Children’s Hospital (IRB–VN01037/IRB00011976/FWA00028418, dated January 15, 2025).

### Authors’ contributions

All authors attest that they meet the ICMJE criteria for authorship. NTT: conception, methodology, data curation, investigation, formal analysis, project administration, writing – original draft; TTTH: conception, methodology, supervision; HBL: methodology, validation, supervision; NVL: data curation, project administration; AWT-R: methodology, supervision, writing – review & editing. All authors: critical review for intellectually important content, approval of manuscript final version, agreement to be accountable for all aspects of the work.

## APPENDIX LABORATORY REFERENCE RANGES

**Supplementary Table 1.** Reference values for blood biochemical analysis.

| Laboratory test | Age | Normal range |
| --- | --- | --- |
| Albumin | 0 – < 15 days | 31 – 43 (g/L) |
|  | 15 days – < 1 year | 28 – 48 |
|  | 1 – < 8 years | 38 – 47 |
|  | 8 – < 15 years | 39 – 49 |
|  | 15 – < 19 years | Female: 38 – 51, Male: 40 – 52 |
| ALT (GPT) | 0 – 5 days | 6 – 50 U/L |
|  | 1 – 19 years | 5 – 40 |
| AST (GOT) | 0 – < 15 days | 30 – 146 (U/L) |
|  | 15 days – < 1 year | 19 – 61 |
|  | 1 – < 7 years | 20 – 40 |
|  | 7 – < 12 years | 17 – 33 |
|  | 12 – < 19 years | Female: 12 – 24, Male: 13 – 32 |
| Total bilirubin | 0 – < 15 days | 4 – 253 (μmol/L) |
|  | 15 days – < 1 year | 2 – 12 |
|  | 1 – < 9 years | 2 – 8 |
|  | 9 – < 12 years | 2 – 10 |
|  | 12 – < 15 years | 3 – 12 |
|  | 15 – < 19 years | 3 – 14 |
| Direct bilirubin | 0 – < 15 days | 3.3 – 7.6 (μmol/L) |
|  | 15 days – < 1 year | 0.1 – 3 |
|  | 1 – < 9 years | 0.1 – 1.8 |
|  | 9 – < 13 years | 0.1 – 2.8 |
|  | 13 – < 19 years | Female: 0.7 – 4, Male: 0.8 – 4.2 |
| CK | Newborn | 468 – 1200 U/L |
|  | ≤ 5 days | 195 – 700 |
|  | < 6 months | 41 – 330 |
|  | > 6 months | 24 – 229 |
| Creatinine | 0 – < 15 days | 30 – 81 μmol/L |
|  | 15 days – < 2 years | 10 – 33 |
|  | 0 – < 5 years | 19 – 38 |
|  | 5 – < 12 years | 28 – 54 |
|  | 12 – < 15 years | 40 – 71 |
|  | 15 – < 19 years | Female: 43 – 73, Male: 55 – 94 |
| CRP | Adult and children | < 6.0 mg/L |
|  | 4 days – 1 month | ≤ 1.6 |
| C3 | 0 – < 15 days | 0.57 – 1.16 g/L |
|  | 15 days – < 1 year | 0.58 – 1.49 |
|  | 1 – < 19 years | 0.85 – 1.42 |
| C4 | 0 – < 1 year | 0.05 – 0.33 g/L |
|  | 1 – < 19 years | 0.12 – 0.41 |
| Ferritin | Newborn | 25 – 200 μg/L |
|  | 1 month | 200 – 600 |
|  | 2 – 5 months | 50 – 200 |
|  | 6 months – 15 years | 7 – 140 |
| K<sup>+</sup> | < 2 months | 3.0 – 6.0 mmol/L |
|  | 2 – 12 months | 3.5 – 5.6 |
|  | > 12 months | 3.5 – 5.0 |
| LDH | 0 – < 15 days | 256 – 1017 U/L |
|  | 15 days – < 1 year | 135 – 375 |
|  | 1 – < 10 years | 159 – 266 |
|  | 10 – < 15 years | 130 – 235 |
|  | 15 – < 19 years | 107 – 207 |
| Na<sup>+</sup> | 0 – 7 days | 133 – 146 mmol/L |
|  | 7 – 31 days | 134 – 144 |
|  | 1 – 6 months | 134 – 142 |
|  | 6 months – 1 year | 133 – 142 |
|  | > 1 year | 134 – 143 |
| Total protein | 1 – 30 days | 41 – 63 g/L |
|  | 31 – 182 days | 44 – 67 |
|  | 183 – 365 days | 55 – 79 |
|  | 1 – 18 years | 57 – 80 |
| Urea | 0 – < 15 days | 1.1 – 8.0 mmol/L |
|  | 15 days – < 1 year | 1.3 – 5.9 |
|  | 1 – < 10 years | 3.2 – 7.7 |
|  | 10 – < 19 years | Female: 2.6 – 6.6, Male: 2.6 – 7.3 |
| Procalcitonin |  | < 0.05 ng/ml |

Alanine aminotransferase (ALT), aka serum glutamic-pyruvic transaminase (GPT); aspartate aminotransferase (AST), aka glutamic-oxaloacetic transaminase (GOT); creatine kinase (CK); C-reactive protein (CRP); potassium (K<sup>+</sup>); lactate dehydrogenase (LDH); sodium (Na<sup>+</sup>)

**Supplementary Table 2.** Reference values for red blood cell analysis.

| Age | RBC (T/L) |  | HGB (g/dL) |  | HCT (%) |  | MCV (fL) |  | MCH (pg) |  | MCHC (g/dL) |  | RDW–CV (%) |  |
| --- | --- | --- | --- | --- | --- | --- | --- | --- | --- | --- | --- | --- | --- | --- |
|  | Male | Female | Male | Female | Male | Female | Male | Female | Male | Female | Male | Female | Male | Female |
| 0 d to < 4 d | 3.69-4.75 | 3.79-4.76 | 12.5-16.6 | 12.7-16.4 | 36.4-47.4 | 36.5-47.7 | 94.0-106.3 | 89.7-105.4 | 32.8-36.4 | 31.7-36.3 | 34.0-35.3 | 33.9-35.4 | 16.3-18.2 | 15.8-17.8 |
| 4 d to < 8 d | 3.98-5.08 | 4.05-4.83 | 12.5-16.3 | 12.6-15.3 | 35.9-46.6 | 36.1-44.0 | 87.1-96.5 | 86.5-93.8 | 30.9-33.4 | 30.6-32.3 | 34.3-35.7 | 34.3-35.7 | 15.0-17.6 | 14.7-16.6 |
| 8 d to < 15 d | 3.75-4.93 | 4.01-4.73 | 11.9-15.7 | 12.7-14.9 | 34.4-45.4 | 36.6-43.2 | 87.1-94.8 | 87.4-92.2 | 30.4-33.0 | 30.5-31.9 | 34.0-35.6 | 33.9-35.3 | 14.9-16.3 | 14.8-16.3 |
| 15 d to < 31 d | 3.61-4.46 | 3.70-4.59 | 11.6-14.2 | 11.6-14.3 | 33.6-41.0 | 34.1-41.8 | 88.0-95.2 | 88.4-93.3 | 30.6-32.6 | 30.5-32.0 | 33.9-35.3 | 33.7-35.1 | 14.6-16.4 | 15.0-16.7 |
| 31 d to < 61 d | 3.24-4.08 | 3.55-4.57 | 10.2-12.7 | 11.1-13.7 | 29.1-36.6 | 32.0-39.9 | 86.5-92.1 | 85.7-91.6 | 30.0-32.0 | 29.8-31.7 | 34.0-35.5 | 34.1-35.4 | 14.7-16.2 | 14.2-15.6 |
| 61 d to < 181 d | 3.67-4.61 | 3.63-4.61 | 10.5-13.0 | 10.7-13.4 | 30.5-37.7 | 30.5-38.6 | 79.6-86.3 | 82.0-87.0 | 27.6-29.9 | 28.5-30.4 | 33.9-35.4 | 34.1-35.6 | 13.5-15.3 | 13.6-14.8 |
| 181 d to < 2 y | 3.81-4.74 | 3.83-4.67 | 10.4-12.5 | 10.8-12.6 | 30.5-36.4 | 30.9-36.4 | 75.6-83.1 | 76.6-83.2 | 26.0-29.0 | 26.5-29.3 | 33.6-35.2 | 34.1-35.6 | 13.6-15.5 | 13.3-14.8 |
| 2 y to < 6 y | 3.92-4.72 | 3.89-4.67 | 11.0-12.8 | 11.1-12.9 | 31.5-36.8 | 31.8-37.0 | 76.8-83.3 | 77.7-84.1 | 26.8-29.4 | 27.0-29.6 | 34.2-35.7 | 34.0-35.6 | 13.2-14.5 | 13.0-14.2 |
| 6 y to < 12 y | 3.85-4.75 | 3.88-4.72 | 11.0-13.3 | 11.3-13.4 | 31.5-38.0 | 32.3-38.3 | 78.2-83.9 | 79.5-85.2 | 27.5-29.7 | 27.8-30.0 | 34.4-35.8 | 34.3-35.8 | 13.0-14.2 | 12.8-13.9 |
| 12 y to < 18 y | 3.74-4.93 | 3.79-4.61 | 11.0-14.3 | 11.3-13.4 | 31.4-41.0 | 32.1-38.7 | 80.8-86.6 | 82.1-87.7 | 28.2-30.5 | 28.4-30.7 | 34.2-35.6 | 33.9-35.4 | 13.0-14.6 | 12.8-14.4 |
| 18 y to adult | 4.54-5.78 | 3.85-5.16 | 13.3-17.2 | 12.0-15.0 | 38.9-50.9 | 34.8-45.0 | 81.2-94.0 | 78.5-96.4 | 27.1-32.5 | 26.4-33.2 | 32.5-36.7 | 31.8-35.9 | 11.5-14.1 | 11.3-14.7 |

RBC – red blood cell; HGB – hemoglobin; HCT – hematocrit; MCV – mean corpuscular volume; MCH – mean corpuscular hemoglobin; MCHC – mean corpuscular hemoglobin concentration; RDW – red blood cell distribution width – coefficient of variation; T/L = tera per liter; g/dL = grams per deciliter; % = percentage; fL = femtoliter (10<sup>-15</sup> L); pg = picogram (10<sup>-12</sup> g)

**Supplementary Table 3.**
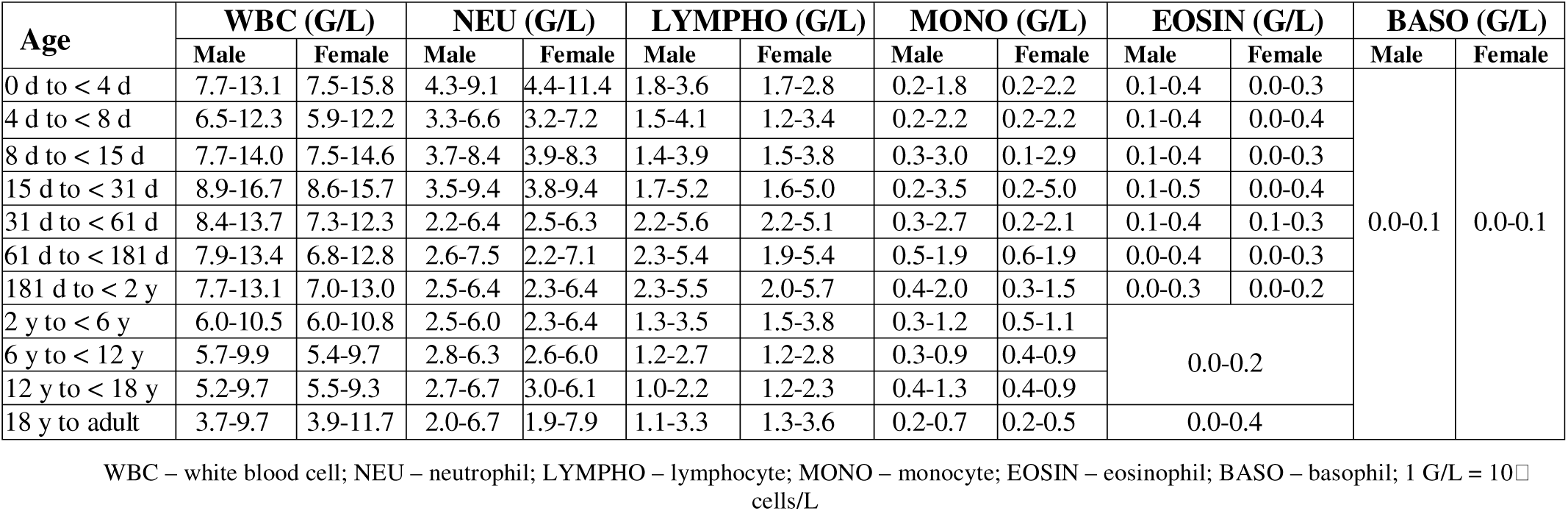
Reference values for white blood cell analysis.

| Age | WBC (G/L) |  | NEU (G/L) |  | LYMPHO (G/L) |  | MONO (G/L) |  | EOSIN (G/L) |  | BASO (G/L) |  |
| --- | --- | --- | --- | --- | --- | --- | --- | --- | --- | --- | --- | --- |
|  | Male | Female | Male | Female | Male | Female | Male | Female | Male | Female | Male | Female |
| 0 d to < 4 d | 7.7-13.1 | 7.5-15.8 | 4.3-9.1 | 4.4-11.4 | 1.8-3.6 | 1.7-2.8 | 0.2-1.8 | 0.2-2.2 | 0.1-0.4 | 0.0-0.3 | 0.0-0.1 | 0.0-0.1 |
| 4 d to < 8 d | 6.5-12.3 | 5.9-12.2 | 3.3-6.6 | 3.2-7.2 | 1.5-4.1 | 1.2-3.4 | 0.2-2.2 | 0.2-2.2 | 0.1-0.4 | 0.0-0.4 |  |  |
| 8 d to < 15 d | 7.7-14.0 | 7.5-14.6 | 3.7-8.4 | 3.9-8.3 | 1.4-3.9 | 1.5-3.8 | 0.3-3.0 | 0.1-2.9 | 0.1-0.4 | 0.0-0.3 |  |  |
| 15 d to < 31 d | 8.9-16.7 | 8.6-15.7 | 3.5-9.4 | 3.8-9.4 | 1.7-5.2 | 1.6-5.0 | 0.2-3.5 | 0.2-5.0 | 0.1-0.5 | 0.0-0.4 |  |  |
| 31 d to < 61 d | 8.4-13.7 | 7.3-12.3 | 2.2-6.4 | 2.5-6.3 | 2.2-5.6 | 2.2-5.1 | 0.3-2.7 | 0.2-2.1 | 0.1-0.4 | 0.1-0.3 |  |  |
| 61 d to < 181 d | 7.9-13.4 | 6.8-12.8 | 2.6-7.5 | 2.2-7.1 | 2.3-5.4 | 1.9-5.4 | 0.5-1.9 | 0.6-1.9 | 0.0-0.4 | 0.0-0.3 |  |  |
| 181 d to < 2 y | 7.7-13.1 | 7.0-13.0 | 2.5-6.4 | 2.3-6.4 | 2.3-5.5 | 2.0-5.7 | 0.4-2.0 | 0.3-1.5 | 0.0-0.3 | 0.0-0.2 |  |  |
| 2 y to < 6 y | 6.0-10.5 | 6.0-10.8 | 2.5-6.0 | 2.3-6.4 | 1.3-3.5 | 1.5-3.8 | 0.3-1.2 | 0.5-1.1 | 0.0-0.2 |  |  |  |
| 6 y to < 12 y | 5.7-9.9 | 5.4-9.7 | 2.8-6.3 | 2.6-6.0 | 1.2-2.7 | 1.2-2.8 | 0.3-0.9 | 0.4-0.9 |  |  |  |  |
| 12 y to < 18 y | 5.2-9.7 | 5.5-9.3 | 2.7-6.7 | 3.0-6.1 | 1.0-2.2 | 1.2-2.3 | 0.4-1.3 | 0.4-0.9 |  |  |  |  |
| 18 y to adult | 3.7-9.7 | 3.9-11.7 | 2.0-6.7 | 1.9-7.9 | 1.1-3.3 | 1.3-3.6 | 0.2-0.7 | 0.2-0.5 | 0.0-0.4 |  |  |  |

WBC – white blood cell; NEU – neutrophil; LYMPHO – lymphocyte; MONO – monocyte; EOSIN – eosinophil; BASO – basophil; 1 G/L = 10<sup>9</sup> cells/L

**Supplementary Table 4.**
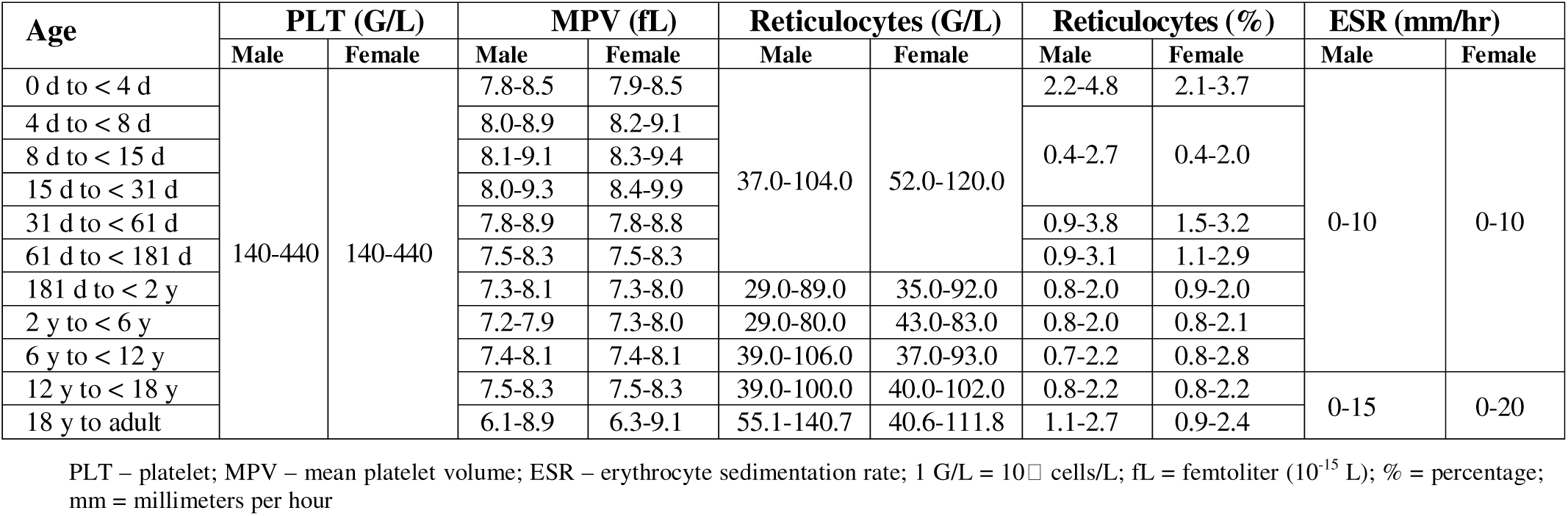
Reference values for further hematological analysis.

**Supplementary Table 5.** Reference values for cerebrospinal fluid analysis.

| Parameter | Age | Normal range |
| --- | --- | --- |
| Total cells (RBC & WBC) | 0 – 1 month | ≤ 27/μl |
|  | 1 month – 16 years: | ≤ 7/μl |
|  | Adult | ≤ 5/μl |
| Total protein | Premature: 27– 32 weeks | 0.68 – 2.4 g/L |
|  | 33 – 36 weeks | 0.67 – 2.3 |
|  | 37 – 40 weeks | 0.58 – 1.5 |
|  | 1 day – 1 month | 0.25 – 0.72 |
|  | 2 – 3 months | 0.20 – 0.72 |
|  | 4 – 6 months | 0.15 – 0.50 |
|  | 7 – 12 months | 0.10 – 0.45 |
|  | 2 years | 0.10 – 0.40 |
|  | 3– 4 years | 0.10 – 0.38 |
|  | 5– 8 years | 0.10 – 0.43 |
|  | Adult | < 0.45 |

567 RBC – red blood cell; WBC – white blood cell; microliter (μl); grams per liter (g/L)

